# Testing for SARS-CoV-2 in care home staff and residents in English care homes: A service evaluation

**DOI:** 10.1101/2020.08.04.20165928

**Authors:** Emma Smith, Clare F Aldus, Julii Brainard, Sharon Dunham, Paul R Hunter, Nicholas Steel, Paul Everden

## Abstract

**Background:** COVID-19 has especially affected care home residents.

**Aim:** To evaluate a nurse-led Enhanced Care Home Team (ECHT) enhanced SARS-CoV-2 testing strategy.

**Design and setting:** Service evaluation in care homes in Norfolk UK.

**Method:** Residents and staff received nose and throat swab tests (7 April to 29 June 2020). Resident test results were linked with symptoms on days 0-14 after test and mortality to 13 July 2020.

**Results:** Residents (n=518) in 44 homes and staff (n=340) in 10 care homes were tested. SARS-CoV-2 positivity was identified in 103 residents in 14 homes and 49 staff in seven homes. Of 103 SARS-CoV-2+ residents, just 38 had typical symptom(s) at time of test (new cough and/or fever). Amongst 54 residents who were completely asymptomatic when tested, 12 (22%) developed symptoms within 14 days. Compared to SARS-CoV-2 negative residents, SARS-CoV-2+ residents were more likely to exhibit typical symptoms (new cough (n=26, p=0.001); fever (n=24, p=<0.001)) or as ‘generally unwell’ (n=18, p=0.001). Of 38 resident deaths, 21 (55%) were initially attributed to SARS-CoV-2, all of whom tested SARS-CoV-2+. One death not initially attributed to SARS-CoV-2 also tested positive.

**Conclusion:** Testing identified asymptomatic and pre-symptomatic SARS-CoV-2+ residents and staff. Being ‘generally unwell’ was common amongst symptomatic residents and may indicate SARS-CoV-2 infection in older people in the absence of more ‘typical’ symptoms. Where a resident appears generally unwell SARS-CoV-2-infection should be suspected. Protocols for testing involved integrated health and social care teams.

## Introduction

Older people residing in care homes are extremely vulnerable to SARS-CoV-2 infection. ^1 2^ Transmission of SARS-CoV-2 may be possible up to two-days prior to the appearance of typical symptoms yet older patients frequently have atypical presentation, ^3-5^ making recognition and control of infection in care homes difficult. Coupled with this, care homes in the UK have been consistently under-resourced, ^6^ staff are largely unregistered, ^7^ and training and support for healthcare support workers is limited. ^7,8^ Carers commonly work across settings on casual contracts. ^9^ Allocation of SARS-CoV-2-tests and personal protective equipment (PPE) supply was initially focussed on clinical settings in the UK.

The British National Health Service (NHS) has a low *per capita* inpatient bed base (compared to other high income countries) which means pressure is high to discharge vulnerable patients to social care settings. ^10,11^ To protect NHS bed-capacity, patients were moved from hospitals to care homes untested until 16 April. ^12^ A national strategy to support formal testing of symptomatic residents and care home workers started from 15 April 2020. Whole care home testing in affected homes started from 15 May 2020, ^12^ and voluntary screening from 11 June 2020. SARS-CoV-2 has highlighted serious gaps in data intelligence surrounding care homes, ^13^ with regional test results typically not available to local authorities until 2 July 2020.^12^

The county of Norfolk lies in the East of England, UK. North Norfolk is one of seven local authority districts within Norfolk and has a total population of approximately 200,000. It has devolved administration for many public services including primary care (North Norfolk Primary Care; NNPC).^14^ North Norfolk has the oldest median age, at 53.8 years, of any local authority area in England and Wales. ^15^ This compares with median age of 45 years across Norfolk, and 40.2 years for the UK. ^15^ Of the 89 registered residential homes for the elderly in North Norfolk, 57 receive enhanced nursing care services (as described below) from NNPC.

In the UK approximately 13.7% of people aged 85 and over live in care homes. ^16^ Care home residents typically have high levels of healthcare needs related to chronic progressive disease including dementia, multiple disabilities and high dependency. Older people and staff of care homes in North Norfolk benefit from a series of well-developed integrated care services including an Enhanced Care Home Team (ECHT) service commissioned by Norfolk and Waveney Clinical Commissioning Group (CCG) in December 2018 and provided by North Norfolk Primary Care (NNPC).^17^ ECHT comprises five nurses (two advanced nurse practitioners (ANP), three nurse practitioners (NP)) and a paramedic. With NNPC GPs, ECHT provides holistic care through consistent GP review of the mental and physical health of their care home patients. Their objectives are to reduce unplanned hospital admissions or readmissions, undertake medicine reviews, and provide palliative care enabling residents to die with dignity and compassion at ‘home’. ECHT works with care homes to identify at-risk patients through risk stratification.

ECHT developed an ‘intelligent’ SARS-CoV-2 testing strategy. At a time when test capacity was low, this prevented unnecessary use of tests yet optimised infection control and care by focussed testing of residents and staff of SARS-CoV-2-affected homes.

## Method

Residents receiving at least one SARS-CoV-2 test result and staff test results are included. In homes where SARS-CoV-2-was identified, all residents and staff were offered screening tests. Broader screening was introduced from 11 June 2020. Resident test results were linked with symptoms recorded in care records at day-0 (test date) to day-14, and mortality outcomes.

### Recruitment

All 57 care homes for which North Norfolk Primary Care were responsible were eligible for ECHT COVID-support. Of these, 44 requested at least one resident SARS-CoV-2-test through ECHT and 10 had staff tested. SARS-CoV-2 testing commenced on 7 April 2020. The results described here cover the monitoring period from 7 April to 29 June 2020.

### SARS-CoV-2 testing

Where SARS-CoV-2-was suspected the resident was isolated and barrier nursing implemented. The testing procedure is described elsewhere. ^18^ Swabs were taken at the home by ANPs (for residents) or at drive-through sites or self-administered (for staff). Analyses (rRT-PCR), conducted locally (Norfolk and Norwich University Hospital Foundation Trust) were reported via electronic medical records (EMR). Where SARS-CoV-2-infection was confirmed, barrier nursing was continued with escalation of care where appropriate.

Staff or residents could receive more than one test. Staff who tested positive were asked to self-isolate for 7-days. Retests were conducted for residents for whom SARS-CoV-2 results were negative if they began or continued to exhibit putative symptoms.

### Clinical presentation

Residents of UK care homes are registered with local primary care services and clinical conditions recorded in EMRs. For SARS-CoV-2-positive residents, data on typical SARS-CoV-2 symptoms (new cough, temperature, anosmia), and atypical symptoms (anorexia, generally unwell, confused or agitated, fatigue, GI disturbance, rash, falls) or any mention of any other symptom (other) at the time of testing were extracted from medical records by an ANP. In addition, for SARS-CoV-2-positive residents who were asymptomatic at the point of test, data on any symptoms recorded in the 14-day post-test period were extracted.

### Mortality

Cause of death data were obtained from residents medical records and death certificates.

### Data analysis

Data were pseudonymised and provided to the investigators by ECHT. Analyses were conducted using STATA (StataCorp v. 16). Data for residents and staff tests comprised unique ID, care home ID, date of SARS-CoV-2 test(s) and test outcome(s). Data for residents also included age, sex and symptoms (SARS-CoV-2-positive residents only).

Residents’ results were reported for individuals but staff test results were reported by home ID. Residents who ever had a SARS-CoV-2-positive test were considered SARS-CoV-2-positive. The number of potential residents per home was based on the number of care home beds.

Estimates of the number of staff tested was based on care home manager report of the proportion of staff tested, the number of staff employed in each home, and number of tests reported.

Cases were asymptomatic if they had no symptoms at the point of test or in the subsequent 14-day period. Cases were presymptomatic if they had no symptoms at the point of test but developed symptoms in the subsequent 14-day period.

SARS-CoV-2 was accepted as the cause of death where SARS-CoV-2 was certified as a cause of death.

## Results

Figure 1 describes testing and screening in the study.

SARS-CoV-2 results for residents (n=518) of 44 care homes who received one or more SARS-CoV-2 test and staff tests (n=545) across 10 care homes in North Norfolk were included. In homes where screening was adopted, 461 of 708 (65.1%) potential residents and an estimated 340 of 434 potential staff (78.3%) were tested. Residents received 618 tests (mean 1.2, range 1 to 4) each and staff 545 (mean 1.6; range unknown) tests each.

**Figure 1.**
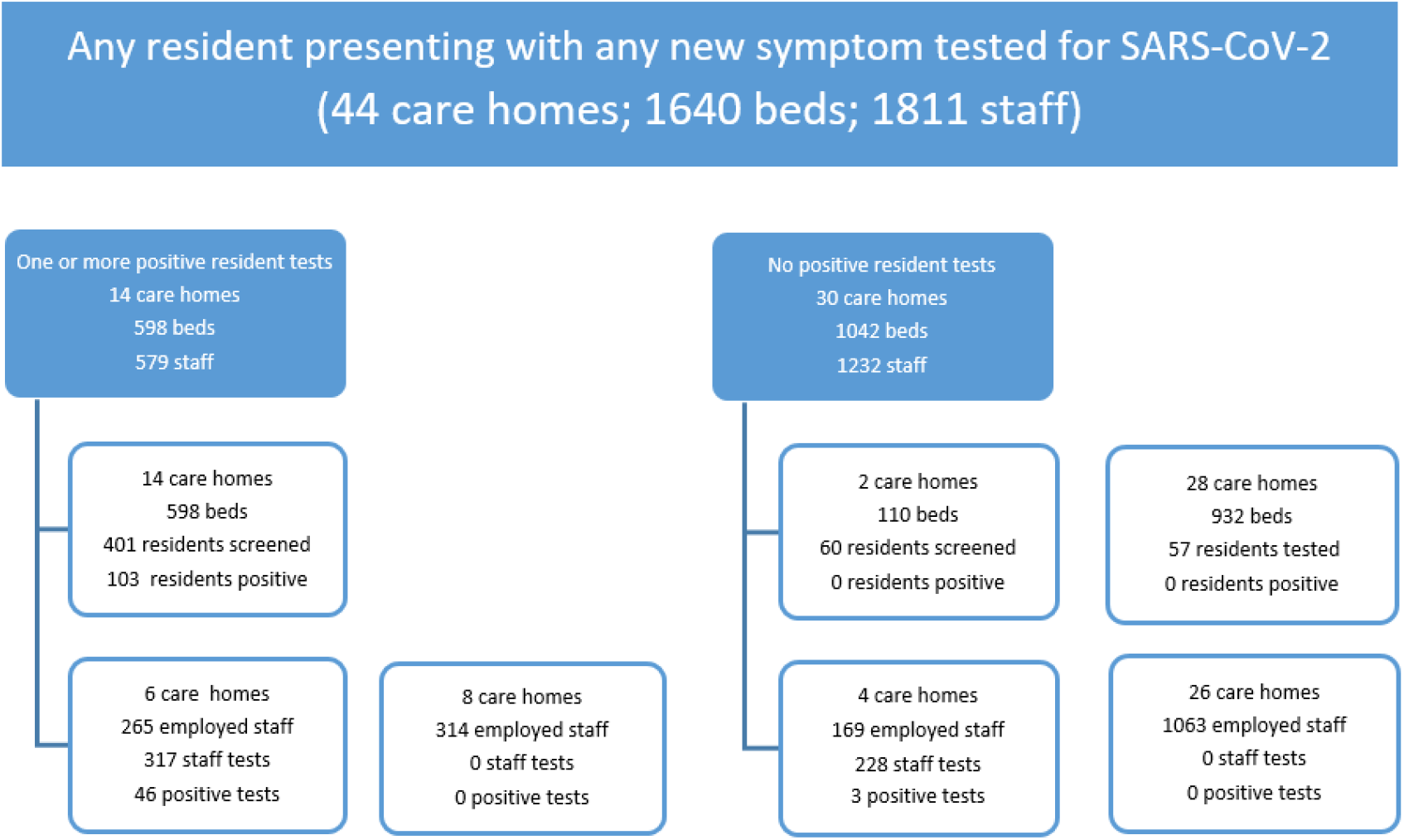
Flow of testing and screening for residents and staff across care homes

Table 1 describes demographic and clinical presentations of residents by SARS-CoV-2 test outcome. The mean age of tested residents was 86.8 years (SD 9.6, range 42-104). More residents were female (n=364, 70%) and females were older (mean 87.6 years; Student’s t-test P=0.0013).

**Table 1.** Demographic and clinical presentation for residents by SARS-CoV-2 test outcome and SARS-CoV-2 test outcomes for staff

|  | Residents(n=518) |  |  | Staff (estimated* n=340) |  |
| --- | --- | --- | --- | --- | --- |
|  | Resident tests (n=618) |  |  | Staff tests (n=545) |  |
| Characteristic | -ve result<br>(%/range) | +ve result<br>(%/range) | P-value | -ve result<br>(%) | +ve result<br>(%) |
| Overall (people) | 415 (80.1) | 103 (19.9) |  | 291 (85.6) | 49 (14.4) |
| Overall (tests) | 515 (83.3) | 103 (16.7) |  | 496 (91.0) | 49 (9.0) |
| Female | 302 (72.7) | 62 (60.1) |  | . | . |
| Age | 86.5 (42-104) | 87.8 (71-104) | 0.237 <sup>S</sup> | . | . |
| Symptomatic | 108 (26.0) | 49 (47.6) |  | . | . |
| CV-typical symptoms | 62 (14.9) | 38 (36.9) |  | . | . |
| Asymptomatic | n/a | 42 (40.8) |  | . | . |
| Pre-symptomatic | n/a | 12 (11.7) |  | . | . |
| New cough | 52 (11.8) | 26 (25.2) | 0.001 <sup>P</sup> | . | . |
| Fever | 25 (6.0) | 24 (23.3) | 0.000 <sup>P</sup> | . | . |
| Anosmia | 0 | 0 | . | . | . |
| Anorexia | 9 (2.2) | 6 (5.8) | 0.048 <sup>P</sup> | . | . |
| Confused/agitated | 14 (3.4) | 6 (5.8) | 0.248 <sup>F</sup> | . | . |
| Falls | 2 (0.5) | 4 (3.9) | 0.016 <sup>F</sup> | . | . |
| Fatigue | 7 (1.7) | 5 (4.9) | 0.069 <sup>F</sup> | . | . |
| GI Disturbance | 14 (3.4) | 5 (4.9) | 0.556 <sup>F</sup> | . | . |
| Generally unwell | 29 (7.0) | 18 (17.5) | 0.001 <sup>P</sup> | . | . |
| Rash | 2 (0.5) | 2 (1.9) | 0.178 <sup>F</sup> | . | . |
| Other (not specified) | 12 (2.9) | 1 (1.0) | 0.480 <sup>F</sup> | . | . |

Symptomatic, any mention of any symptom; Asymptomatic, no symptoms at test date or during the subsequent 14-day period; Pre-symptomatic, asymptomatic at test date but any mention of any symptoms in subsequent 14-day period. <sup>P</sup>, Pearson's $\chi^2$ ; <sup>F</sup>, Fisher's exact test; <sup>S</sup>, Student's T-test. \*Estimates of the number of staff tested were based on care home manager report of the proportion of staff tested, the number of staff employed in each home, and number of tests reported.

### SARS-CoV-2 test results

Most homes (n=30, 68%) had no SARS-CoV-2-infections. In 14 care homes where SARS-CoV-2 was identified, 103 (25.6% residents tested; 17.2% beds) and 49 (14.4% staff (estimated); 9% of staff tests) were SARS-CoV-2-positive.

Where both staff and residents were tested, there was close correlation between positive resident and staff groups with SARS-CoV-2-positive staff and resident groups in 6 homes and SARS-CoV-2-negative staff and residents in three. In one care home, three staff but no patients were SARS-CoV-2-positive.

SARS-CoV-2-positive residents were similar in age to SARS-CoV-2-negative residents (p=0.237) and more likely to be male (Pearson’s χ2, p=0.012).

### Clinical presentation

Of the 103 SARS-CoV-2-positive residents 49 (47.6%) had any symptoms and 38 (36.9%) had typical SARS-CoV-2 symptoms (new cough or fever).

At point of test, 54 (52.4%) SARS-CoV-2-positive residents had no symptoms. Of these, 42 (40.8%) remained asymptomatic during the following 14 days. The remaining 12 (11.6%) were pre-symptomatic and developed one or more symptoms during the following 14-days.

Clinical presentations are shown (Table 1) by SARS-CoV-2-status. SARS-CoV-2-positive residents were more likely to exhibit typical symptoms (new cough, n=26, p=0.001; fever, n=24, p=<0.001) than SARS-CoV-2-negative residents. Further, SARS-CoV-2-positive residents were more likely to present as ‘generally unwell’ (n=18; p=0.001) than SARS-CoV-2-negative residents.

Typical symptoms of SARS-CoV-2 are new cough, fever, and anosmia. ^19^ New cough was identified in 26 (25.2%) residents, fever in 24 (23.3%). Anosmia was not detected, but within this cohort may be undiagnostic because around 62.5% people aged 80- to 97 have olfactory impairment and self-reported olfactory impairment in older people is low. ^20^

### Mortality

Table 2 describes the distribution of SARS-CoV-2 cases, test outcomes and deaths by care home. 38 deaths were recorded. Death attributed to SARS-CoV-2 (n=21, 55%) occurred across eight homes. Non-SARS-CoV-2 deaths (n=17, 45%) included dementia (n=7); old age or expected death (n=3); multi-organ failure (n=1); bronchopneumonia (n=1); intracranial haematoma (n=1) and unknown causes (n=4). All deaths attributed to SARS-CoV-2 tested positive for SARS-CoV-2. One of 17 deaths not attributed to SARS-Co-V-2 was positive for SARS-CoV-2.

**Table 2.** SARS-CoV-2 test outcomes by residents and staff and resident mortality by care home.

| Home ID | SARS-CoV-2 test result resident |  |  | SARS-CoV-2 test result staff* |  |  | SARS-CoV-2 attributed-resident-death |  |  |
| --- | --- | --- | --- | --- | --- | --- | --- | --- | --- |
|  | N | Pos | % | N* | Pos | % | N | Pos | % |
| 1 | 8 | 0 | 0 |  |  |  |  |  |  |
| 2 | 4 | 0 | 0 |  |  |  |  |  |  |
| 3 | 20 | 1 | 5 | 99 | 4 | 4 |  |  |  |
| 4 | 62 | 13 | 21 | 70 | 9 | 13 | 3 | 2 | 67 |
| 5 | 9 | 1 | 11 |  |  |  |  |  |  |
| 6 | 1 | 0 | 0 |  |  |  |  |  |  |
| 7 | 2 | 0 | 0 |  |  |  |  |  |  |
| 8 | 34 | 21 | 62 | 63 | 19 | 30 | 4 | 3 | 75 |
| 9 | 59 | 18 | 31 |  |  |  | 5 | 4 | 80 |
| 10 | 13 | 12 | 92 | 7 | 1 | 14 | 1 | 1 | 100 |
| 11 | 21 | 1 | 5 |  |  |  | 2 | 1 | 50 |
| 12 | 25 | 0 | 0 | 162 | 3 | 2 | 1 |  | 0 |
| 13 | 37 | 9 | 24 | 20 | 2 | 10 | 12 | 6 | 50 |
| 14 | 5 | 0 | 0 |  |  |  |  |  |  |
| 15 | 2 | 0 | 0 |  |  |  | 1 | 0 | 0 |
| 16 | 1 | 0 | 0 |  |  |  |  |  |  |
| 17 | 1 | 0 | 0 |  |  |  |  |  |  |
| 18 | 4 | 0 | 0 |  |  |  | 1 | 0 | 0 |
| 19 | 42 | 3 | 7 |  |  |  | 1 | 0 | 0 |
| 20 | 35 | 0 | 0 |  |  |  | 2 | 0 | 0 |
| 21 | 9 | 1 | 11 |  |  |  |  |  |  |
| 22 | 28 | 16 | 57 |  |  |  | 2 | 2 | 100 |
| 23 | 3 | 0 | 0 |  |  |  |  |  |  |
| 24 | 2 | 0 | 0 |  |  |  |  |  |  |
| 25 | 1 | 0 | 0 |  |  |  |  |  |  |
| 26 | 16 | 2 | 13 | 58 | 11 | 19 |  |  |  |
| 27 | 34 | 3 | 9 |  |  |  | 2 | 2 | 100 |
| 28 | 1 | 0 | 0 | 25 | 0 | 0 |  |  |  |
| 29 | 1 | 0 | 0 |  |  |  |  |  |  |
| 30 | 1 | 0 | 0 | 21 | 0 | 0 |  |  |  |
| 31 | 3 | 0 | 0 | 20 | 0 | 0 |  |  |  |
| 32 | 17 | 2 | 12 |  |  |  |  |  |  |
| 33 | 1 | 0 | 0 |  |  |  |  |  |  |
| 34 | 1 | 0 | 0 |  |  |  |  |  |  |
| 35 | 3 | 0 | 0 |  |  |  |  |  |  |
| 36 | 2 | 0 | 0 |  |  |  |  |  |  |
| 37 | 1 | 0 | 0 |  |  |  |  |  |  |
| 38 | 1 | 0 | 0 |  |  |  |  |  |  |
| 39 | 1 | 0 | 0 |  |  |  |  |  |  |
| 40 | 1 | 0 | 0 |  |  |  |  |  |  |
| 41 | 1 | 0 | 0 |  |  |  |  |  |  |
| 42 | 2 | 0 | 0 |  |  |  |  |  |  |
| 43 | 1 | 0 | 0 |  |  |  |  |  |  |
| 44 | 2 | 0 | 0 |  |  |  | 1 | 0 | 0 |
| **Total** | **518** | **103** | **20** | **545** | **49** | **9** | **38** | **21** | **55** |

Note: \*all staff-tests in each care home

## Discussion

### Summary

For staff and residents who were tested under this study around one in six residents and one in seven staff, were SARS-CoV-2-positive. In 10 homes where staff and residents were tested there was close correlation of SARS-CoV-2-infection. SARS-CoV-2-positive staff and resident groups were identified in six homes and SARS-CoV-2-negative staff and residents in three, illustrating the binary nature of infection in care homes: i.e., either the care home community has infections or it does not.

Importantly, amongst atypical presentations it was common for residents to be ‘generally unwell’. There is a risk that physical symptoms from SARS-CoV-2 infection may go unrecognised particularly due to mental health co-morbidity. ^21^ Identification of SARS-CoV-2 in older people should not be based solely upon the presence of typical symptoms. Until vaccination is available, residents presenting with any new symptom or who are ‘generally unwell’ should be considered putative SARS-CoV-2 cases.

Overall 54 (52.4%) SARS-CoV-2-positive residents were asymptomatic at the point of test.

Of these, 42 (78%) did not subsequently develop symptoms (true asymptomatic) and 12 (22%) developed symptoms (pre-symptomatic). The high proportions of asymptomatic or pre-symptomatic cases underline the value of screening to break chains of transmission.

### Strengths and limitations

ECHT were able to rapidly develop and implement early SARS-CoV-2-testing. Early screening of residents and staff after ingress into care homes identified prevalence of truly asymptomatic infections and symptom presentation in residents relatively early in the UK COVID-19 outbreak.

Staff results were reported by home (not individual) and therefore numbers of staff tested are estimated. Prevalence of staff infection and the possible relationship between staff prevalence and resident prevalence could be explored. The potential value of better information on staff working practices (knowing who works in other settings) was evidenced. In addition, resident estimates are based on bed number. Bed capacity is close to but not consistently at 100%. Many residents may not be tested for ethical or clinical reasons.

### Comparison with existing literature

The Vivaldi study, a telephone survey of care home managers exploring whole-home testing across 9,081 care homes in England (26 May to 19 June 2020), found 20% of residents and 7% staff SARS-CoV-2-positive in homes where infection was reported. ^22^ Our findings for residents in screened homes are broadly consistent with Vivaldi (17.2% versus 20%). However, this study identified higher prevalence of infection amongst staff compared to Vivaldi (14.4 versus 7%). This may be because Vivaldi was based on care home manager report and this study on screening. Graham *et* al.; identified 40% SARS-CoV-2-infection amongst residents in four care homes where serious outbreaks were already in progress. ^23^ The comparatively low proportion (17.2% vs. 40%) amongst ECHT-study residents of affected homes may be due to early identification and isolation of cases in Norfolk.

Infection was identified in 14 of 44 (32%) EHCT-study care homes. Vivaldi reported infection in 56% of care homes. ^22^ Comparatively low prevalence of affected ECHT-study care homes may be due to the low prevalence of infection in Norfolk. Care homes in most regions had a lower chance of infection compared to care homes in London. ^22^

Staff but no residents were SARS-CoV-2-positive in one study care home. This is consistent with the findings of McNichol et al. ^24^ Vivaldi identified that staff members increase the odds of infection for residents by 11%,^22^ further underlining the importance of screening staff.

Cross-setting working was noted during our study. The Vivaldi study identified that around 12% of care homes have staff that work across settings which increases the odds (OR 2.40, 95% confidence interval: 1.92 to 3.00) of infection in staff compared to care homes who have staff who never work elsewhere. ^22^ Reduction in transmission of infection between care homes may be supported by understanding across-setting working. No routinely collected data currently capture multi-setting working in North Norfolk.

### Clinical presentation

True asymptomatic cases were also reported from the Diamond Princess cruise ship (17.9%), ^25^ Japanese citizens evacuated from Wuhan (30.8%), ^26^ and in a care facility for older people in the USA (6.3%).^27^ A comparable US study of 11 care homes identified 55.4% (n=507) asymptomatic cases. ^28^ World Health Organisation advice is that transmission from asymptomatic persons is less likely than from symptomatic people. ^29^

### Mortality

Under this study, 21 of 38 (55%) deaths were attributed to SARS-CoV-2. In the period 10 April 2020 to 26 June 2020 the UK Care Quality Commission (CQC) reported 12,211 (34.8%) and 133 (24.5%) all-cause deaths attributed to SARS-CoV-2 of residents of care homes in England and Norfolk respectively. ^30^ Over the period 10 April 2020 to 29 May 2020, Carterwood *et al*. estimated that Norfolk had 84 fewer deaths than expected given the local prevalence of SARS-CoV-2.^31^ A high proportion of deaths (21/38, 55%) attributed to SARS-CoV-2 infection in this study is expected because screening was typically conducted in homes where infection was confirmed. Mortality data for care homes in North Norfolk cannot be disaggregated from other areas of Norfolk so it is not possible to deduce whether the ECHT intervention reduced mortality.

### Implications for research and practice

Enhanced SARS-CoV-2-testing and screening enabled identification of SARS-CoV-2-related deaths that might otherwise have gone unrecognised. SARS-CoV-2 screening in care homes supported accurate attribution of mortality.

Asymptomatic or atypical presentation is common amongst SARS-CoV-2-positive care home residents. Where a resident appears generally unwell or has any new symptom, SARS-CoV-2-infection should be suspected. Where SARS-CoV-2-infection is found, residents and staff should be screened. Early testing and screening of staff and residents in care homes can accurately identify outbreaks, prevalence of infection and death, and cause of death. Integrated health and social care teams working closely with care homes are well-placed to implement rapid screening services. Protocols for early screening which include local integrated health and social care teams should be developed. Comparative evaluation of this service was difficult because relevant data were non-existent: we did not know which staff work across settings or how many infections or deaths had occurred in neighbouring care homes. Integrated health and social care datasets that support urgent and local service development and evaluation should be commissioned.

### How this fits in

Spread of COVID-19 can be reduced by early detection and monitoring regimes in residential care homes for the elderly, but how to best achieve early disease detection in these settings remains unclear. Understanding both typical and atypical symptom prevalence in both residents and staff may be vital to breaking chains of transmission. Early in the UK COVID-19 outbreak, an integrated nursing support team was able to quickly implement a testing regime that thus documented a range of presentations among both residents and staff and especially helped to identify residents who were pre-symptomatic or asymptomatic.

## Data Availability

Data are available on request from the corresponding author.

## Acknowledgements

We thank the staff and residents of study care homes who took part in this evaluation.

## Ethical Approval

Approvals for this service evaluation were granted to authors PE and CA by Norfolk and Waveney Clinical Commissioning Group (5 June 2020) and University of East Anglia Faculty of Medicine and Health Ethics Committee (15 June 2020, reference 2019/20-139), respectively.

## Authors’ contributions

PE conceived, developed and led implementation of the ECHT-led North Norfolk SARS-CoV-2 care home testing service; ES and SD extracted resident and staff data from patient health records and care homes; CA conducted analyses; and ES, CA, JB, NS, PH, PE developed the manuscript. All authors have read and agree to the published version of the manuscript.

